# Modifiable risk factors and inflammation-related proteins in polymyalgia rheumatica: genome-wide meta-analysis and Mendelian randomisation

**DOI:** 10.1101/2024.04.21.24306135

**Authors:** Sizheng Steven Zhao, Sarah L Mackie, Susanna C Larsson, Stephen Burgess, Shuai Yuan

## Abstract

**Objective:** Polymyalgia rheumatica (PMR) is an age-related inflammatory disease of unknown cause. We aimed to identify potentially modifiable risk factors and therapeutic targets for preventing or treating PMR.

**Methods:** We meta-analysed genetic association data from 8,156 cases of PMR (defined using diagnostic codes and self-report) and 416,495 controls of European ancestry from the UK Biobank and FinnGen. We then performed Mendelian randomization analyses to estimate the association between eight modifiable risk factors (using data from up to 1.2 million individuals) and 65 inflammation-related circulating proteins (up to 55,792 individuals), using the inverse variance weighted and pleiotropy robust methods.

**Results:** We identified three novel genome-wide significant loci in the *IL1R1, NEK6* and *CCDC88B* genes and confirmation of previously described associations with *HLA-DRB1* and *ANKRD55*. Genetically predicted smoking intensity (OR 1.32; 95%CI 1.08-1.60; p=0.006) and visceral adiposity (OR 1.22; 95%CI 1.10-1.37; p=3.10x10^−4^) were associated with PMR susceptibility. Multiple circulating proteins related to IL-1 family signaling were associated with PMR. IL-1 receptor-like 2, also known as IL-36 receptor (OR 1.25; p=1.89x10^−32^), serum amyloid A2 (OR 1.06, 9.91x10^−10^) and CXCL6 (OR 1.09, p=4.85x10^−7^) retained significance after correction for multiple testing.

**Conclusion:** Reducing smoking and visceral adiposity at a population level might reduce incidence of PMR. We identified proteins that may play causal roles in PMR, potentially suggesting new therapeutic opportunities. Further research is needed before these findings are applied to clinical practice.

**Key messages:**

1. Three novel PMR risk loci were identified in *IL1R1, NEK6* and *CCDC88B*.
2. Several circulating proteins, notably those involved in IL-1 family signalling, are associated with PMR susceptibility.
3. Visceral adiposity and cigarette smoking are causally associated with risk of PMR.

## Introduction

Polymyalgia rheumatica (PMR) is an age-related inflammatory disease of unknown cause (1). The pattern of inflammation of PMR centres on certain tendon, ligaments, and joint capsules, but without radiographic change in adjacent articular bone (2). There is overlap with giant cell arteritis (GCA) and both diseases have been postulated to preferentially initiated at mechanically-stressed sites (3,4). The intense systemic inflammatory response of PMR hampers efforts to disentangle true causal factors from epiphenomena related to systemic inflammation or compensatory/counter-regulatory mechanisms. To date, poor understanding of PMR aetiopathogenesis has been a barrier to progress in its definition, diagnosis, and treatment. Currently, long-term oral glucocorticoids are the primary treatment used, at the cost of substantial treatment toxicity (1).

Like GCA, PMR is more commonly diagnosed in northern European countries, but genetic and environmental contribution to this geographical distribution is unclear. Genome-wide association studies have yielded mechanistic insights into GCA implicating angiogenesis and neutrophils (5). Historically, genetic association data in PMR has been contradictory or inconclusive due to underpowering and historic use of candidate gene methods (1). Recently, we reported that genetically proxied IL-6 receptor (6) and IL-1 receptor antagonism (7) are both associated with reduced risk of PMR. In the current genetic investigation, we aimed to identify potentially modifiable risk factors and inflammation-related circulating proteins as targets of intervention for preventing or treating PMR.

## Methods

### Study design

We performed a genome wide association study (GWAS) meta-analysis using data from the UK Biobank and FinnGen. Genetic association data was used in Mendelian randomization (MR) analyses of selected modifiable risk factors and inflammation-related circulating proteins. Ethical approval was obtained by the FinnGen and UK Biobank (application 72723) studies to which participants provided written informed consent.

### GWAS of UK Biobank and FinnGen

The UK Biobank is a cohort study of approximately 500,000 individuals aged 40-69 years, recruited between 2006 and 2010 (8). It includes self-reported data on diseases, linked hospital admissions data, the national death register, and primary care data for a subset of ∼225,000 participants. Quality control of UK Biobank genotype data has been described previously (8). The current analysis was restricted to unrelated (kinship≥0.084) individuals of self-reported White-British ancestry, with concordant submitted and genetically inferred sex. GWAS was performed using SNPTEST v2.5.4, adjusting for the first four principal components derived using cases and controls. Variants with minor allele frequency of <0.01 and INFO score of <0.5 were excluded. PMR was defined using any of the following: ICD 10 code M353 (from fields 41270, 40001 or 40002), self-report (code 1377 in field 20002), or Read code N20.. or XE1FJ in linked primary care data. GWAS comprised 4,285 cases of PMR (3180 ICD, 1073 self-report, 1239 Read) and 17,140 randomly selected controls without PMR (1:4 ratio to reduce case-control imbalance that can increase type I error (9,10)) and was adjusted for the first four principal components.

FinnGen is a large research study comprising genetic and health data from 500,000 Finnish participants. Quality control steps have been previously described. We used Release 10 (December 2023) summary-level data on PMR, comprising 3,871 cases and 399,355 controls. PMR was defined by ICD code 10th (M353), 9th (725), or 8th (4463) revisions. Genetic associations were adjusted for age, sex, principal components, and genotyping batch.

We combined summary statistics from UK Biobank and FinnGen under a fixed effects model using the METAL software (11) and examined the heterogeneity in SNP-PMR associations across the two studies. LD Score regression was used to evaluate the heritability, genomic inflation factor (lambda), and intercept (12).

### Mendelian randomization analysis

MR analyses used genetic association data from the above GWAS meta-analysis comprising a total of 8,156 PMR cases and 416,495 controls. We first investigated the association between eight modifiable environmental or lifestyle risk factors and PMR, including three obesity indicators (body mass index (BMI) (13), waist circumference (14), and visceral adiposity (15)) and five lifestyle factors (smoking initiation (16), smoking intensity measured in cigarettes per day (16), alcohol (16) and coffee consumption (17), and moderate-to-vigorous physical activity (18)). To identify potential therapeutic targets, we conducted MR analyses of 66 inflammation-related circulating proteins (19–21) in relation to PMR susceptibility. **Table S1** provides information on GWASs from which genetic association data for these exposures were derived.

For the eight modifiable risk factors, we selected SNPs associated with the exposures of interest at the genome-wide significance threshold (*P*⍰<⍰5×10^−8^) from their respective GWASs. For the inflammation-related circulating proteins, only *cis*-acting SNPs (i.e., within ±250 kb of the gene encoding the relevant protein) were selected. For all exposures, we excluded SNPs exhibiting high linkage disequilibrium (LD) (*r*^*2*^ > 0.01, using the 1000 Genomes European reference panel), retaining only the SNP with the lowest *P* value from the GWAS association. For inflammation-related circulating proteins with a limited number of SNPs as genetic instruments, we sought proxy SNPs that had a high LD (*r*^*2*^ > 0.8) with those SNPs that were not available.

For exposures with two SNPs as instruments, we used the inverse variance weighted method under a fixed effects model to estimate their association with risk of PMR; otherwise, we used the multiplicative random effects inverse variance weighted method. For exposures proxied by multiple SNPs, we conducted sensitivity analyses, including the weighted median and MR-Egger methods, to validate the robustness of our findings and detect and address potential horizontal pleiotropy. We used Cochran’s Q value to assess the heterogeneity among the SNP estimates. Multiple testing corrections were implemented using the Benjamini-Hochberg false discovery rate (FDR) method. All analyses were based on two-sided tests and performed using the TwoSampleMR and MendelianRandomization packages in R (version 4.1.1).

## Results

### Genome-wide association analysis

This GWAS meta-analysis included 424,651 individuals (8,156 cases and 416,495 non-cases). Assuming a lifetime prevalence of 2% among those of European ancestry (22), the heritability of PMR on a liability scale explained by common genetic variants (minor allele frequency (MAF) ≥0.05) was estimated at *h*^*2*^⍰= 0.09 (95% CI 0.06-0.11). The genomic inflation factor, λ_GC_, was calculated to be 1.06, suggesting a minimal inflation due to population stratification or other confounding factors. The LD score regression intercept was estimated at 0.99 (standard error=0.008), indicating a good alignment with the null expectation of no polygenicity.

We annotated five genome-wide significant loci (**Figure 1**), with *IL1R1* (rs11694915), *NEK6* (rs748741), *CCDC88B* (rs615540) being previously unreported (**Table 1**), and *HLA-DRB1* and *ANKRD55* consistent with prior reports (23). rs2760985 (tagging *HLA-DRB1*) was the most pronounced association observed. It was also the only SNP with statistical evidence of heterogeneity across FinnGen and UK Biobank cohorts, but estimates were directionally concordant (**Table S2**).

**Table 1.** Genetic loci associated with PMR at the genome-wide significance level.

| rsID for lead variant | Variant position (Ch37/hg19) | Nearest gene | EA | NEA | EAF | Beta | SE | *P value* | OR | Previously known |
| --- | --- | --- | --- | --- | --- | --- | --- | --- | --- | --- |
| rs11694915 | 2:102710068 | *IL1R1* | A | T | 0.3865 | -0.0978 | 0.0172 | 1.40E-08 | 0.91 | No |
| rs748741 | 9:127089775 | *NEK6* | A | G | 0.6168 | -0.0959 | 0.0172 | 2.31E-08 | 0.91 | No |
| rs615540 | 11:64123838 | *CCDC88B* | A | G | 0.5489 | 0.1056 | 0.0169 | 3.70E-10 | 1.11 | No |
| rs2760985 | 6:32566398 | *HLA-DRB1* | A | G | 0.2181 | 0.5307 | 0.0209 | 7.78E-142 | 1.70 | Yes |
| rs7731626 | 5:55444683 | *ANKRD55* | A | G | 0.3123 | -0.1618 | 0.0181 | 4.59E-19 | 0.85 | Yes |

EA, effect allele; EAF, effect allele frequency; NEA, non-effect allele; SE, standard error.

**Figure 1.**
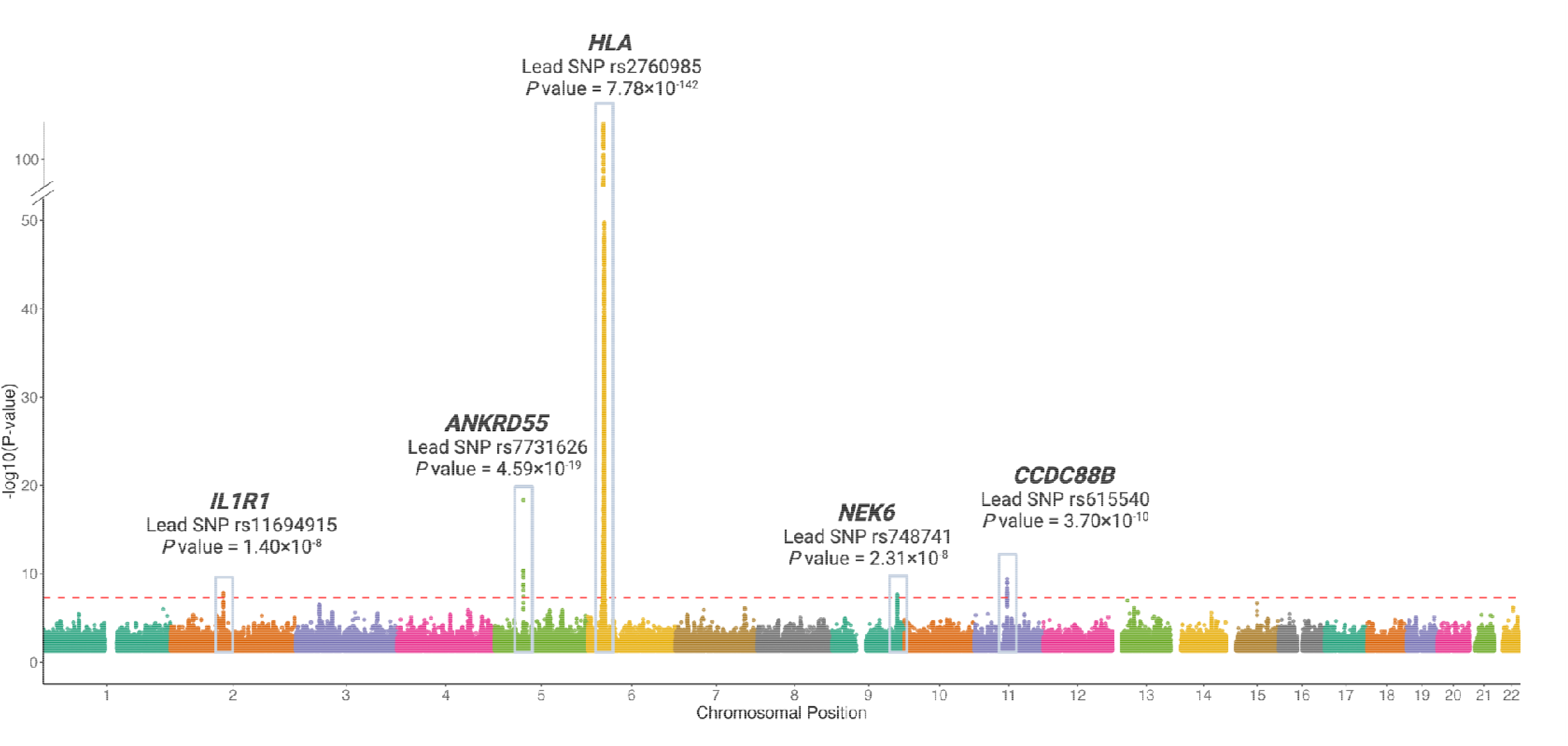
Results of genome-wide association study in PMR and summary of 5 identified genomic risk loci. SNPs, single nucleotide polymorphisms.

### Mendelian randomization analysis

Of the eight modifiable risk factors, genetically predicted levels of BMI, visceral adiposity and smoking intensity (cigarettes per day) were associated with PMR (**Figure 2**). Genetically predicted smoking intensity was associated with 1.32-fold higher odds of PMR per standard deviation (SD) increase (95%CI 1.08-1.60; p=0.006). Higher genetically predicted BMI (OR 1.16 per SD increase; 95%CI 1.03-1.31; p=0.013) and visceral adiposity (OR 1.22 per SD increase; 95%CI 1.10-1.37; p=3.10×10^−4^) were also associated with PMR risk. After false discovery rate (FDR) multiple-testing correction, associations with visceral adiposity and smoking intensity retained significance. There was no statistical evidence of directional pleiotropy (**Table S3**). Genetically predicted levels of the other modifiable risk factors showed no significant associations with PMR susceptibility.

**Figure 2.**
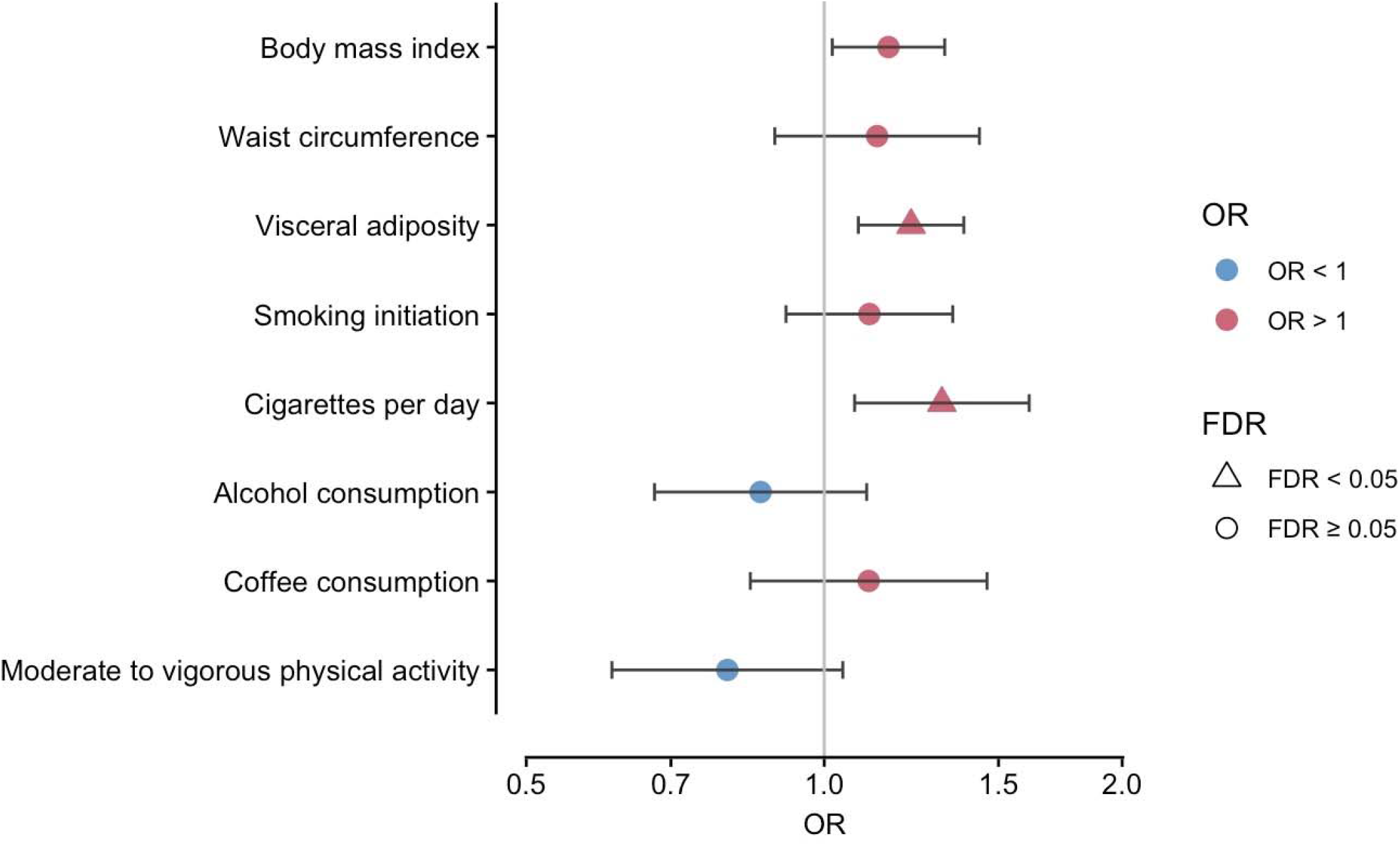
Associations between genetically proxied modifiable factors and the risk of PMR in Mendelian randomization analysis. FDR, false discovery rate correction for eight risk factors. Bars show 95% confidence interval. OR, odds ratio, estimate per standard deviation increase in all risk factors, except for coffee consumption (per 50% change in intake) and physical activity (being active verses inactive).

Of the inflammation-related circulating proteins, suitable genetic instruments were identified for 65. Genetically predicted levels of 12 inflammatory markers were associated with susceptibility to PMR (**Figure 3**) including various proteins in the IL-1 family signaling pathways. After adjusting for multiple testing, only the associations for IL-1 receptor-like 2 (also known as IL-36 receptor) (OR 1.25; 95%CI 1.21, 1.30; p=1.89x10^−32^), serum amyloid A2 protein (OR 1.06; 95%CI 1.04, 1.07; p=9.91x10^−10^) and CXCL6 (CXC motif chemokine 6) (OR 1.09; 95%CI 1.05, 1.13; p=4.85x10^−7^) remained significant. Full results for all inflammation-related circulating proteins are shown in **Table S4**. The three SNPs used to instrument IL-1 receptor-like 2 are in close proximity to the *IL1R1* SNP identified in GWAS and may represent the same underlying association; rs3917265 is correlated with rs11694915 (*IL1R1*) with r^2^ of 0.11).

**Figure 3.**
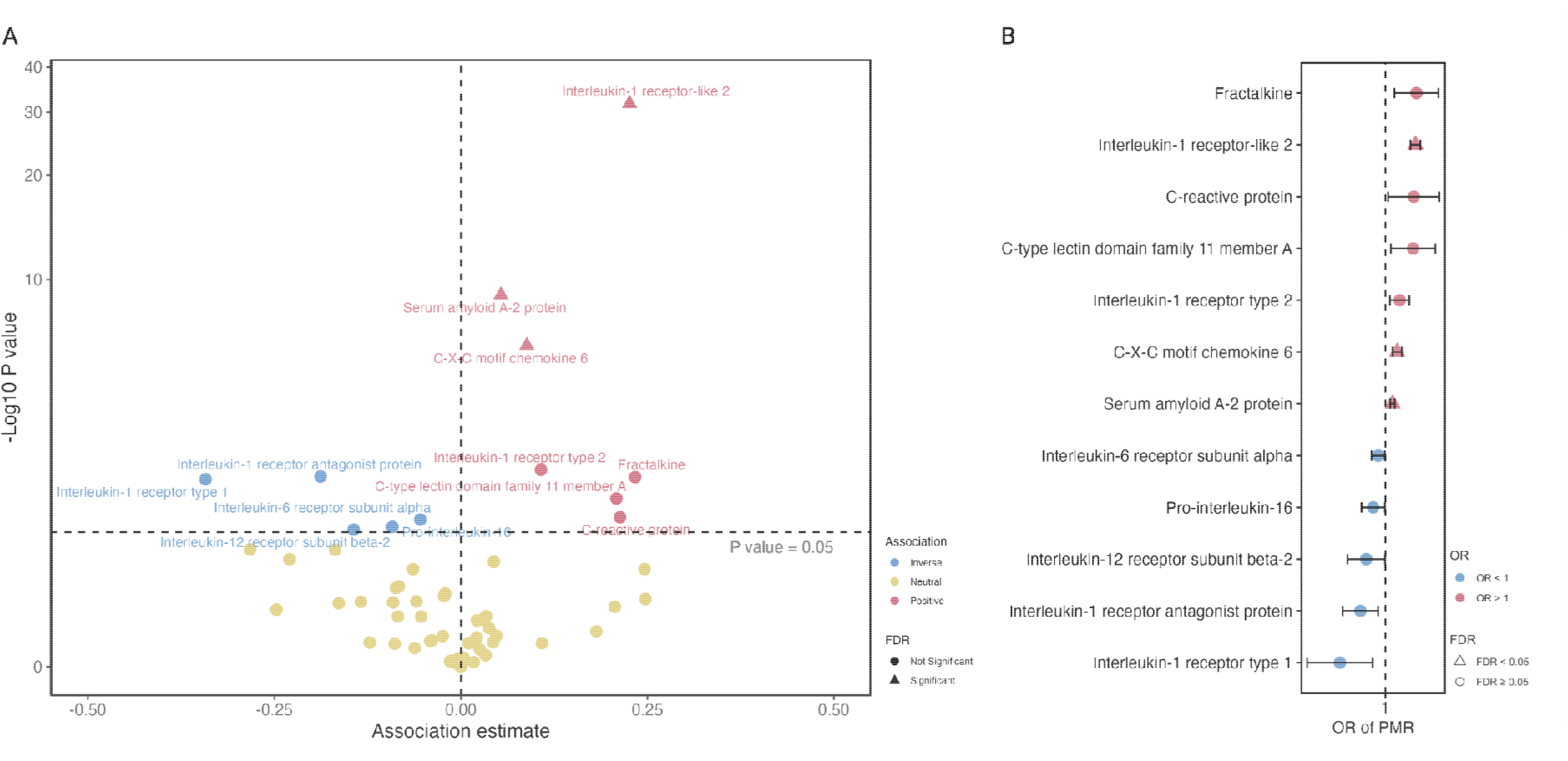
Associations between genetically predicted levels of inflammatory markers and the risk of PMR in Mendelian randomization analysis. A. volcano plot of MR associations between 65 inflammatory markers and PMR. B. forest plot of 12 associations with P value < 0.05. Estimates were scaled to one standard deviation increase in genetically predicted markers.

## Discussion

This, the largest GWAS meta-analysis of PMR to date, has identified three novel loci in the *IL1R1, NEK6* and *CCDC88B* genes and confirmation of previously described associations with *HLA-DRB1* and *ANKRD55*. Moreover, we found causal associations between PMR and levels of circulating proteins involved in IL-1 family signalling (potentially explained by the same IL-1 GWAS signal), the acute phase response (serum amyloid A2), and neutrophil chemotaxis, angiogenesis and tendon repair (CXCL6). MR analyses identified visceral adiposity and cigarette smoking as modifiable risk factors for PMR.

The next largest published genetic association study of PMR, to our knowledge, was an exome-wide analysis of 559 cases and 15,620 controls (24). That prior study identified two SNPs in the HLA region, rs3096702 (which the authors could not replicate) upstream of *Notch4* and rs6910071 in *C6orf10*. A conference abstract that similarly combined UK Biobank and an earlier release of FinnGen data identified SNPs close to the *HLA-DRB1* gene and the same intronic variant in *ANKRD55* as in the current study (23). *ANKRD55* variants have been linked to expression of the neighbouring *IL6ST* gene in various immune cells (25). *IL6ST* encodes part of a cytokine receptor complex that act as a signal transducer shared by multiple cytokines, including IL-6. These findings are supported by a prior Mendelian randomization study linking genetically proxied IL-6R inhibition and reduced PMR risk (6), and additionally by the current results for IL-6 receptor subunit alpha. These genetic findings may be viewed as supporting the relevance of IL-6 signalling in PMR, consistent with clinical trial efficacy of IL-6R inhibition (26,27).

Genes encoding proteins involved in IL-1 signalling have been identified in prior candidate gene studies of PMR, for example *IL1RN* (28). We previously showed that genetically proxied IL-1R antagonism is associated with PMR risk (7). Multiple IL-1 family members were implicated in the current hypothesis-free genome-wide and inflammatory-protein-wide association analyses, but most did not survive correction for multiple testing. Canakinumab, an IL-1b inhibitor, reduced PMR disease activity in a 2-week proof of concept study of five patients (29) but large-scale trials of IL-1 antagonism in PMR are lacking. The only IL-1 family protein to survive multiple testing was IL-1 receptor like 2, also known as the IL-36 receptor. Activation of the IL-36 receptor can induce trained immunity in monocytes (30) and has a variety of other functions (31) depending on cellular context. IL-36 receptor inhibitors have been approved for generalised pustular psoriasis (32) but not yet trialled in PMR.

Serum amyloid A2, one of the inducible serum amyloid A (SAA) proteins, was the only hepatic acute-phase protein that survived multiple testing in our analysis. In addition to being highly inducible in liver by IL-6 and IL-1, this lipophilic protein also acts as a damage-associated molecular pattern (DAMP) molecule and may be secreted by macrophages or adipocytes (33,34). *In vitro*, SAA can induce macrophage IL-1b secretion via activation of the NLRP3 inflammasome; in the circulation this effect is abrogated by SAA binding to high-density lipoprotein, which inactivates it (35). SAA can also recruit neutrophils and activate their glycolytic pathways *in vitro* (36). In an *ex vivo* model of GCA serum amyloid A can promote angiogenesis (37). Serum amyloid A1 and A2 have also been implicated in other Th17-associated inflammatory diseases (38). A phase III trial of secukinumab for PMR is ongoing (NCT04930094).

CXCL6, also known as GCP-2, is one of the pro-angiogenic chemokines, originally described as a neutrophil chemoattractant. In macrophage and endothelial cell lines, SAA synergises with CRP to upregulate CXCL6 alongside CXCL8, another neutrophil chemoattractant (39). Fibroblasts from patients with ulcerative colitis produce IL-6 upon stimulation by IL-17 (40). In a study that used single-cell techniques to characterise cell subpopulations from healthy and diseased human tendons taken from near the myotendinous junction, CXCL6 was among the inflammatory/alarmin genes that were overexpressed by microfibril-associated tenocytes in chronic tendinopathy (41). Intriguingly, in a model of tendon injury in the horse, IL-1b, released after injury, “licensed” mesenchymal stem cells (MSCs) to secrete an array of reparative and immunomodulatory factors, including CXCL6 (42). In the mouse meniscus-defect model, extracellular vesicles (EVs) secreted by human MSCs upregulated local chondrocyte and MSC CXCL5 and CXCL6 expression; the EVs promoted chondrocyte growth and migration and this effect could be abrogated by blocking CXCL5 and CXCL6, or their common receptor. It was concluded that CXCL5 and CXCL6 were involved in stimulating cartilage response to injury (43). CXCL6 is a multifunctional molecule. Via its action on (human) CXCR1/2, it promotes cartilage repair (44); but also, via binding to local glycosaminoglycans, CXCL6 creates a chemokine concentration gradient that brings in neutrophils that can exacerbate tissue damage (45).

Challenges of interpreting observational studies illustrate the complex relationship between adiposity, PMR susceptibility and glucocorticoid toxicity. On the one hand, active (undiagnosed) PMR causes weight loss. IL-6 has a key role in thermoregulation; it is produced by muscle in response to exercise, promoting lipolysis, and is also secreted by beige adipocytes in response to cold exposure. IL-6 itself promotes further “beiging” to convert pro-inflammatory white adipose tissue towards thermogenic “beige” adipose tissue (46), promoting energy utilisation and weight loss. On the other hand, glucocorticoid treatment causes visceral adiposity, which is strongly correlated with cardiovascular risk. Aging can also lead to accumulation of visceral adipose tissue. Similarly, smoking can alter both innate and adaptive immune responses, with persistent effects especially on adaptive immunity long after individuals quit smoking (47) as well as impairing tissue perfusion, healing and angiogenesis. It remains to be seen whether smoking and adiposity, as well as being amenable to lifestyle change, might also prove useful for identifying patients with PMR who are at higher risk of adverse outcomes.

This study highlights previously-unappreciated aspects of the aetiopathogenesis of PMR, a disease with a characteristic pattern of inflammation of certain tendon and ligament sites (2) that are under high mechanical stress (4). In young individuals, mechanical stress from physical activity or high BMI elicits in adaptive changes such as increase in cross-sectional area and tendon stiffness; however, this ability to respond to mechanical stress is lost in older people (48). Visceral adipose tissue contains activated immune cells producing IL-1b and IL-6 with systemic effects. Fat deposition in and around muscles, entheses and tendons occurs with aging or obesity, and may contribute to chronic tendinopathy (49).

Antagonistic pleiotropy is a theory to explain why genes causing age-associated diseases can persist in populations despite natural selection. Hypothetically, a genetic tendency to greater IL-1 induced local CXCL6 release might hasten healing of injuries in youth, and protect joints from damage; but delayed healing in aging, or the pro-inflammatory effect of visceral adiposity, would lead to a chronic damage-repair cycle with persistent local inflammation and neoangiogenesis, that might be exacerbated by the same genetic tendency. This hypothesis could be further investigated in healthy volunteers of different genotypes, ages and visceral adiposity levels.

The comparatively large number of PMR cases is a strength of the current study, but there are also limitations. First, our analyses focused on populations of European ancestry, potentially limiting the generalizability of our findings to other populations. Second, PMR cases were defined using clinical diagnostic codes, which may introduce misclassification. There is a lack of genetic data from large-scale inception cohorts of well-characterised PMR patients. Furthermore, there are no formal diagnostic criteria for PMR and thus no gold standard other than expert clinical opinion. In practice, the clinical diagnostic boundaries between PMR and “elderly-onset” seronegative RA are defined by clinical convention and consensus. Better mechanistic understanding of the differences between these two diseases, and between PMR and GCA, may enable dividing-lines between these diseases to be drawn more accurately in future. Multimodal approaches including genetic evidence will be needed.

In conclusion, this study identified three novel PMR risk loci in *IL1R1, NEK6* and *CCDC88B* and confirmed previously described associations with *HLA-DRB1* and *ANKRD55*. The importance of the *IL1R1* locus was supported by causal associations between circulating proteins involved in IL-1 family signalling and PMR risk. Results also suggested visceral adiposity and cigarette smoking to be potentially modifiable risk factors for PMR. These findings improve the current understanding of the genetic underpinning of PMR, and propose potential new avenues for prevention and treatment.

## Acknowledgements

SSZ is supported by a National Institute for Health Research (NIHR) Academic Clinical Lectureship. SLM is supported in part by the NIHR Leeds Biomedical Research Centre (grant NIHR203331). The views expressed are those of the authors and not necessarily those of the National Health Service, the NIHR, or the Department of Health. This work was supported by Versus Arthritis (grant number 21173, grant number 21754 and grant number 21755) and by the NIHR Manchester Biomedical Research Centre. SB is supported by the Wellcome Trust (225790/Z/22/Z) and the United Kingdom Research and Innovation Medical Research Council (MC_UU_00002/7, MC_UU_00040/01).

## Data availability statement

UK Biobank data are available to all bona fide researchers for use in health-related research that is in the public interest. The application procedure is described at www.ukbiobank.ac.uk. GWAS meta-analysis data will be made publicly available upon publication.

## Ethics approval

Ethical approval was obtained by the UK Biobank study. The current analysis was performed under application number 72723.

## Patient and Public Involvement

Patients or the public were not involved in the design, or conduct, or reporting, or dissemination plans of this research.

