## supplementary materials for "Modifiable risk factors and inflammation-related proteins in polymyalgia rheumatica: genome-wide meta-analysis and Mendelian randomisation"

.

### Table S1. Details for genome-wide association studies from which data were obtained.

| Exposure | No. SNPs | *F* statistic | Unit | PubMed ID | Sample size | Ancestry | Measurement |
| --- | --- | --- | --- | --- | --- | --- | --- |
| Body mass index | 311 | 77.2 | SD | 30239722 | 806,834 | European | Calculated as weight (kg) divided by height (m) squared. Weight and height data were measured (most studies) or self-reported. |
| Waist circumference | 573 | 55.8 | SD | 30239722 | 697,734 | European | Measured (most studies) or self- reported. |
| Visceral adiposity | 294 | 50.2 | SD | 31501611 | 397,170 | European | Measured by DXA among 5,109 participants and then predicted among 502,638 using the calibrated prediction models. |
| Smoking initiation | 313 | 40.0 | SD in prevalence of smoking initiation | 36477530 | 2,669,029 | European | Smoking behaviours were measured by questionnaires. Smoking initiation is a binary phenotype representing whether an individual had ever smoked cigarettes regularly in their life. |
| Smoking intensity (cigarettes/day) | 65 | 110.8 | SD in number of cigarettes per day | 36477530 | 618,489 | European | Self-reported average number of cigarettes per day among current and former smokers. |
| Alcohol consumption | 114 | 81.0 | SD increase of log- transformed alcoholic drinks/wk | 36477530 | 2,428,851 | European | Self-reported by questionnaires. |
| Coffee consumption | 12 | 196.1 | 50% change | 31046077 | 375,833 | European | Assessed by a 24-h recall questionnaire. |
| Moderate to vigorous physical activity | 16 | 40.6 | Being active vs. inactive | 36071172 | 298,506 | European | Measured by questionnaires and dichotomized due to the zero- inflated negative binomial nature of the distribution. |
| Inflammatory markers | 1-19 | ≥29.6 | SD | 37205426 | 876-55,792 | European | Measured by BioRad BioPlex assays, SomaScan, or Olink platforms. |

SD, standard deviation; SNPs, single nucleotide polymorphisms.

### Table S2. Genetic associations from the meta-analysis and individual populations.

|  |  | Meta-analysis | | | | | FinnGen | | | | UKB | | | |
| --- | --- | --- | --- | --- | --- | --- | --- | --- | --- | --- | --- | --- | --- | --- |
| SNP | **Chromosome: position** | **EA/OA** | **Beta** | **SE** | **p** | **p_het** | **EAF** | **Beta** | **SE** | **P** | **EAF** | **Beta** | **SE** | **P** |
| rs11694915 | 2:102710068 | A/T | -0.098 | 0.017 | 1.40E-08 | 0.954 | 0.433 | -0.099 | 0.023 | 2.44E-05 | 0.331 | -0.097 | 0.026 | 1.48E-04 |
| rs748741 | 9:127089775 | A/G | -0.096 | 0.017 | 2.31E-08 | 0.694 | 0.379 | 0.102 | 0.024 | 1.49E-05 | 0.388 | 0.089 | 0.025 | 3.77E-04 |
| rs615540 | 11:64123838 | A/G | 0.106 | 0.017 | 3.70E-10 | 0.171 | 0.471 | -0.084 | 0.023 | 3.13E-04 | 0.429 | -0.130 | 0.025 | 1.12E-07 |
| rs2760985 | 6:32566398 | A/G | 0.531 | 0.021 | 7.78E-142 | 0.0001 | 0.150 | 0.453 | 0.029 | 3.83E-54 | 0.290 | 0.613 | 0.030 | 7.85E-93 |
| rs7731626 | 5:55444683 | A/G | -0.162 | 0.018 | 4.59E-19 | 0.825 | 0.279 | -0.166 | 0.026 | 2.95E-10 | 0.342 | -0.158 | 0.025 | 2.61E-10 |

EA, effect allele; OA, other allele; SE standard error; p_het, p-value for heterogeneity; EAF, effect allele frequency; UKB, UK Biobank.

### Table S3. Mendelian randomization estimates for the association between risk factors and polymyalgia rheumatica.

|  |  | IVW-fixed | | | IVW-random | | | | Weighted median | | | | MR-Egger | | | | |
| --- | --- | --- | --- | --- | --- | --- | --- | --- | --- | --- | --- | --- | --- | --- | --- | --- | --- |
| Exposure | **No. SNP** | **beta** | **se** | **p** | **beta** | **se** | **p** | **beta** | | **se** | **p** | **beta** | | **se** | **p** | **intercept** | **intercept p** |
| Body mass index | 311 | 0.149 | 0.060 | 0.013 | 0.149 | 0.067 | 0.025 | 0.142 | | 0.116 | 0.221 | 0.251 | | 0.164 | 0.127 | -0.002 | 0.499 |
| Waist circumference | 43 | 0.123 | 0.096 | 0.202 | 0.123 | 0.121 | 0.311 | 0.252 | | 0.163 | 0.121 | 0.590 | | 0.324 | 0.076 | -0.015 | 0.128 |
| Visceral adiposity | 294 | 0.202 | 0.056 | 0.0003 | 0.202 | 0.063 | 0.001 | 0.223 | | 0.103 | 0.030 | 0.181 | | 0.200 | 0.368 | 0.000 | 0.910 |
| Smoking initiation | 346 | 0.105 | 0.085 | 0.218 | 0.105 | 0.099 | 0.288 | 0.117 | | 0.131 | 0.371 | 0.925 | | 0.405 | 0.023 | -0.010 | 0.037 |
| Cigarettes per day | 65 | 0.274 | 0.099 | 0.006 | 0.274 | 0.104 | 0.008 | 0.198 | | 0.159 | 0.214 | 0.299 | | 0.192 | 0.125 | -0.001 | 0.878 |
| Alcohol consumption | 114 | -0.148 | 0.126 | 0.241 | -0.148 | 0.126 | 0.240 | -0.072 | | 0.226 | 0.751 | -0.359 | | 0.294 | 0.224 | 0.003 | 0.427 |
| Coffee consumption | 11 | 0.103 | 0.126 | 0.413 | 0.103 | 0.140 | 0.462 | 0.058 | | 0.160 | 0.717 | -0.138 | | 0.270 | 0.621 | 0.013 | 0.322 |
| Moderate to vigorous physical activity | 17 | -0.225 | 0.181 | 0.214 | -0.225 | 0.137 | 0.100 | -0.109 | | 0.243 | 0.655 | 0.370 | | 0.971 | 0.709 | -0.015 | 0.542 |

### Table S4. Mendelian randomization estimates for the association between inflammatory markers and polymyalgia rheumatica.

|  |  |  | IVW-fixed | | | IVW-random | | | Weighted median | | | MR-Egger | | | | |
| --- | --- | --- | --- | --- | --- | --- | --- | --- | --- | --- | --- | --- | --- | --- | --- | --- |
| Exposure | **No. SNP** | **FDR** | **beta** | **se** | **p** | **beta** | **se** | **p** | **beta** | **se** | **p** | **beta** | **se** | **p** | **intercept** | **intercept p** |
| Interleukin-1 receptor-like 2 | 3 | 1.23E-30 | 0.226 | 0.063 | 3.09E-04 | 0.226 | 0.019 | 1.89E-32 | 0.233 | 0.066 | 4.31E-04 | 0.222 | 0.180 | 0.434 | 0.001 | 0.984 |
| Serum amyloid A-2 protein | 3 | 3.22E-08 | 0.054 | 0.044 | 2.26E-01 | 0.054 | 0.009 | 9.91E-10 | 0.052 | 0.046 | 2.51E-01 | 0.050 | 0.088 | 0.673 | 0.002 | 0.966 |
| C-X-C motif chemokine 6 | 4 | 1.05E-05 | 0.088 | 0.038 | 2.06E-02 | 0.088 | 0.018 | 4.85E-07 | 0.090 | 0.038 | 1.76E-02 | 0.542 | 0.570 | 0.443 | -0.188 | 0.509 |
| Interleukin-1 receptor type 2 | 2 | 5.96E-02 | 0.107 | 0.037 | 4.10E-03 | 0.107 | 0.136 | 4.29E-01 |  |  |  |  |  |  |  |  |
| Fractalkine | 1 | 5.96E-02 | 0.234 | 0.085 | 5.80E-03 |  |  |  |  |  |  |  |  |  |  |  |
| Interleukin-1 receptor type 1 | 2 | 5.96E-02 | -0.342 | 0.126 | 6.42E-03 | -0.342 | 0.293 | 2.44E-01 |  |  |  |  |  |  |  |  |
| Interleukin-1 receptor antagonist protein | 4 | 5.96E-02 | -0.188 | 0.085 | 2.64E-02 | -0.188 | 0.068 | 5.63E-03 | -0.219 | 0.093 | 1.92E-02 | -0.348 | 0.357 | 0.432 | 0.027 | 0.690 |
| C-type lectin domain family 11 member A | 1 | 1.19E-01 | 0.208 | 0.085 | 1.46E-02 |  |  |  |  |  |  |  |  |  |  |  |
| Interleukin-6 receptor subunit alpha | 11 | 2.11E-01 | -0.054 | 0.025 | 2.90E-02 | -0.054 | 0.025 | 3.25E-02 | -0.040 | 0.035 | 2.56E-01 | -0.115 | 0.099 | 0.274 | 0.026 | 0.540 |
| C-reactive protein | 1 | 2.11E-01 | 0.213 | 0.098 | 3.00E-02 |  |  |  |  |  |  |  |  |  |  |  |
| Pro-interleukin-16 | 5 | 2.47E-01 | -0.092 | 0.046 | 4.47E-02 | -0.092 | 0.045 | 4.18E-02 | -0.084 | 0.050 | 9.26E-02 | -0.032 | 0.061 | 0.641 | -0.022 | 0.238 |
| Interleukin-12 receptor subunit beta-2 | 1 | 2.50E-01 | -0.144 | 0.072 | 4.61E-02 |  |  |  |  |  |  |  |  |  |  |  |
| E-selectin | 1 | 4.00E-01 | -0.283 | 0.164 | 8.56E-02 |  |  |  |  |  |  |  |  |  |  |  |
| C-X-C motif chemokine 16 | 2 | 4.00E-01 | -0.168 | 0.098 | 8.62E-02 | -0.168 | 0.034 | 4.96E-07 |  |  |  |  |  |  |  |  |
| Placenta growth factor | 1 | 4.91E-01 | -0.230 | 0.145 | 1.13E-01 |  |  |  |  |  |  |  |  |  |  |  |
| Interleukin-1 receptor-like 1 | 17 | 4.94E-01 | 0.044 | 0.028 | 1.15E-01 | 0.044 | 0.028 | 1.22E-01 | 0.036 | 0.038 | 3.39E-01 | -0.014 | 0.113 | 0.904 | 0.026 | 0.602 |
| Interleukin-2 receptor subunit beta | 1 | 5.35E-01 | 0.246 | 0.170 | 1.48E-01 |  |  |  |  |  |  |  |  |  |  |  |
| C-C motif chemokine 7 | 1 | 5.35E-01 | -0.064 | 0.044 | 1.48E-01 |  |  |  |  |  |  |  |  |  |  |  |
| Interleukin-17D | 1 | 7.66E-01 | -0.087 | 0.073 | 2.36E-01 |  |  |  |  |  |  |  |  |  |  |  |
| C-C motif chemokine 4 | 5 | 7.66E-01 | -0.083 | 0.078 | 2.88E-01 | -0.083 | 0.069 | 2.26E-01 | -0.127 | 0.097 | 1.87E-01 | -0.230 | 0.144 | 0.209 | 0.038 | 0.312 |
| C-C motif chemokine 8 | 1 | 7.67E-01 | -0.021 | 0.019 | 2.74E-01 |  |  |  |  |  |  |  |  |  |  |  |
| Haptoglobin | 17 | 7.67E-01 | -0.021 | 0.019 | 2.77E-01 | -0.021 | 0.019 | 2.67E-01 | -0.010 | 0.026 | 7.12E-01 | 0.073 | 0.063 | 0.269 | -0.056 | 0.141 |
| C-C motif chemokine 16 | 1 | 7.67E-01 | -0.023 | 0.021 | 2.83E-01 |  |  |  |  |  |  |  |  |  |  |  |
| Fibroblast growth factor 23 | 1 | 7.67E-01 | 0.247 | 0.239 | 3.02E-01 |  |  |  |  |  |  |  |  |  |  |  |
| Pro-epidermal growth factor | 1 | 7.67E-01 | -0.134 | 0.135 | 3.20E-01 |  |  |  |  |  |  |  |  |  |  |  |
| C-X-C motif chemokine 5 | 1 | 7.67E-01 | -0.091 | 0.092 | 3.25E-01 |  |  |  |  |  |  |  |  |  |  |  |
| Hepatocyte growth factor | 1 | 7.67E-01 | -0.164 | 0.168 | 3.30E-01 |  |  |  |  |  |  |  |  |  |  |  |
| Growth-regulated alpha protein | 5 | 7.67E-01 | -0.060 | 0.069 | 3.86E-01 | -0.060 | 0.060 | 3.18E-01 | -0.099 | 0.076 | 1.93E-01 | -0.215 | 0.115 | 0.158 | 0.050 | 0.190 |
| Eotaxin | 1 | 7.98E-01 | 0.206 | 0.224 | 3.56E-01 |  |  |  |  |  |  |  |  |  |  |  |
| Interleukin-2 receptor subunit alpha | 1 | 8.17E-01 | -0.247 | 0.280 | 3.77E-01 |  |  |  |  |  |  |  |  |  |  |  |
| Vascular endothelial growth factor A | 11 | 8.52E-01 | -0.053 | 0.058 | 3.57E-01 | -0.053 | 0.068 | 4.33E-01 | -0.065 | 0.080 | 4.15E-01 | 0.143 | 0.121 | 0.266 | -0.039 | 0.093 |
| Serum amyloid A-1 protein | 5 | 8.52E-01 | 0.034 | 0.039 | 3.80E-01 | 0.034 | 0.043 | 4.30E-01 | 0.049 | 0.041 | 2.27E-01 | 0.066 | 0.079 | 0.467 | -0.015 | 0.650 |
| C-C motif chemokine 27 | 1 | 8.52E-01 | -0.084 | 0.107 | 4.32E-01 |  |  |  |  |  |  |  |  |  |  |  |
| C-C motif chemokine 14 | 4 | 8.65E-01 | 0.027 | 0.027 | 3.03E-01 | 0.027 | 0.037 | 4.60E-01 | 0.032 | 0.030 | 2.83E-01 | 0.036 | 0.137 | 0.816 | -0.007 | 0.952 |
| C-C motif chemokine 3 | 3 | 8.65E-01 | 0.022 | 0.046 | 6.40E-01 | 0.022 | 0.030 | 4.66E-01 | 0.020 | 0.045 | 6.54E-01 | 0.025 | 0.076 | 0.798 | -0.001 | 0.963 |
| C-C motif chemokine 17 | 3 | 9.54E-01 | 0.048 | 0.057 | 4.01E-01 | 0.048 | 0.094 | 6.15E-01 | 0.068 | 0.070 | 3.32E-01 | 0.069 | 0.475 | 0.908 | -0.006 | 0.970 |
| C-C motif chemokine 22 | 3 | 9.54E-01 | -0.039 | 0.058 | 5.03E-01 | -0.039 | 0.090 | 6.66E-01 | 0.026 | 0.074 | 7.27E-01 | 0.150 | 0.142 | 0.481 | -0.062 | 0.369 |
| Interleukin-23 receptor | 2 | 9.54E-01 | 0.038 | 0.062 | 5.39E-01 | 0.038 | 0.060 | 5.24E-01 |  |  |  |  |  |  |  |  |
| C-C motif chemokine 2 | 1 | 9.54E-01 | 0.182 | 0.319 | 5.69E-01 |  |  |  |  |  |  |  |  |  |  |  |
| Serum amyloid P-component | 2 | 9.54E-01 | -0.024 | 0.049 | 6.19E-01 | -0.024 | 0.002 | 7.65E-46 |  |  |  |  |  |  |  |  |
| Interleukin-17 receptor A | 9 | 9.54E-01 | 0.012 | 0.027 | 6.47E-01 | 0.012 | 0.031 | 6.93E-01 | 0.015 | 0.032 | 6.32E-01 | -0.018 | 0.061 | 0.780 | 0.016 | 0.580 |
| Fibroblast growth factor 7 | 1 | 9.54E-01 | -0.041 | 0.097 | 6.70E-01 |  |  |  |  |  |  |  |  |  |  |  |
| Fibrinogen gamma chain | 2 | 9.54E-01 | 0.043 | 0.107 | 6.87E-01 | 0.043 | 0.145 | 7.66E-01 |  |  |  |  |  |  |  |  |
| C-C motif chemokine 20 | 1 | 9.54E-01 | -0.122 | 0.307 | 6.90E-01 |  |  |  |  |  |  |  |  |  |  |  |
| Antithrombin-III | 1 | 9.54E-01 | 0.109 | 0.279 | 6.97E-01 |  |  |  |  |  |  |  |  |  |  |  |
| Vascular endothelial growth factor C | 1 | 9.54E-01 | 0.010 | 0.026 | 6.99E-01 |  |  |  |  |  |  |  |  |  |  |  |
| Interleukin-8 | 1 | 9.54E-01 | -0.089 | 0.234 | 7.04E-01 |  |  |  |  |  |  |  |  |  |  |  |
| Pro-adrenomedullin | 4 | 9.54E-01 | 0.021 | 0.108 | 8.45E-01 | 0.021 | 0.044 | 6.34E-01 | 0.054 | 0.121 | 6.53E-01 | -0.093 | 0.422 | 0.846 | 0.013 | 0.806 |
| Plasminogen activator inhibitor 1 | 1 | 9.98E-01 | -0.062 | 0.199 | 7.55E-01 |  |  |  |  |  |  |  |  |  |  |  |
| Tumor necrosis factor ligand superfamily member 10 | 2 | 9.98E-01 | 0.025 | 0.088 | 7.73E-01 | 0.025 | 0.160 | 8.74E-01 |  |  |  |  |  |  |  |  |
| Prothrombin | 1 | 9.98E-01 | 0.033 | 0.170 | 8.45E-01 |  |  |  |  |  |  |  |  |  |  |  |
| Interleukin-27 receptor subunit alpha | 1 | 9.98E-01 | 0.003 | 0.023 | 8.78E-01 |  |  |  |  |  |  |  |  |  |  |  |
| C-C motif chemokine 25 | 1 | 9.98E-01 | -0.004 | 0.029 | 8.92E-01 |  |  |  |  |  |  |  |  |  |  |  |
| C-C motif chemokine 5 | 1 | 9.98E-01 | -0.012 | 0.120 | 9.17E-01 |  |  |  |  |  |  |  |  |  |  |  |
| Macrophage colony-stimulating factor 1 | 1 | 9.98E-01 | -0.015 | 0.154 | 9.25E-01 |  |  |  |  |  |  |  |  |  |  |  |
| Interleukin-7 receptor subunit alpha | 1 | 9.98E-01 | -0.009 | 0.105 | 9.33E-01 |  |  |  |  |  |  |  |  |  |  |  |
| Interleukin-18 | 1 | 9.98E-01 | -0.007 | 0.086 | 9.34E-01 |  |  |  |  |  |  |  |  |  |  |  |
| C-X-C motif chemokine 10 | 1 | 9.98E-01 | 0.017 | 0.221 | 9.38E-01 |  |  |  |  |  |  |  |  |  |  |  |
| Macrophage migration inhibitory factor | 1 | 9.98E-01 | -0.007 | 0.111 | 9.49E-01 |  |  |  |  |  |  |  |  |  |  |  |
| C-X-C motif chemokine 11 | 1 | 9.98E-01 | -0.004 | 0.074 | 9.57E-01 |  |  |  |  |  |  |  |  |  |  |  |
| Intercellular adhesion molecule 1 | 3 | 9.98E-01 | -0.003 | 0.059 | 9.59E-01 | -0.003 | 0.051 | 9.53E-01 | 0.018 | 0.065 | 7.81E-01 | 0.084 | 0.094 | 0.535 | -0.026 | 0.447 |
| Mannose-binding protein C | 2 | 9.98E-01 | 0.001 | 0.020 | 9.64E-01 | 0.001 | 0.049 | 9.85E-01 |  |  |  |  |  |  |  |  |
| Interleukin-17 receptor D | 1 | 9.98E-01 | 0.001 | 0.057 | 9.85E-01 |  |  |  |  |  |  |  |  |  |  |  |
| C-X-C motif chemokine 9 | 1 | 9.98E-01 | -0.001 | 0.122 | 9.92E-01 |  |  |  |  |  |  |  |  |  |  |  |
| Interleukin-6 receptor subunit beta | 6 | 9.98E-01 | 0.000 | 0.051 | 9.98E-01 | 0.000 | 0.048 | 9.98E-01 | 0.002 | 0.057 | 9.74E-01 | 0.014 | 0.086 | 0.875 | -0.004 | 0.843 |
